# Robust SARS-CoV-2-specific and heterologous immune responses after natural infection in elderly residents of Long-Term Care Facilities

**DOI:** 10.1101/2021.08.13.21261889

**Authors:** Gokhan Tut, Tara Lancaster, Megan S. Butler, Panagiota Sylla, Eliska Spalkova, David Bone, Nayandeep Kaur, Christopher Bentley, Umayr Amin, Azar T. Jadir, Samuel Hulme, Morenike Ayodel, Alexander C. Dowell, Hayden Pearce, Sandra Margielewska-Davies, Kriti Verma, Samantha Nicol, Jusnara Begum, D. Blakeway, Elizabeth Jinks, Elif Tut, Rachel Bruton, Maria Krutikov, Madhumita Shrotri, Rebecca Giddings, Borscha Azmi, Chris Fuller, Aidan Irwin-Singer, Andrew Hayward, Andrew Copas, Laura Shallcross, Paul Moss

## Abstract

Long term care facilities (LTCF) provide residential and/or nursing care support for frail and elderly people and many have suffered from a high prevalence of SARS-CoV-2 infection. Although mortality rates have been high in LTCF residents there is little information regarding the features of SARS-CoV-2-specific immunity after infection in this setting or how this may influence immunity to other infections. We studied humoral and cellular immunity against SARS-CoV-2 in 152 LTCF staff and 124 residents over a prospective 4-month period shortly after the first wave of infection and related viral serostatus to heterologous immunity to other respiratory viruses and systemic inflammatory markers. LTCF residents developed high levels of antibodies against spike protein and RBD domain which were stable over 4 months of follow up. Nucleocapsid-specific responses were also elevated in elderly donors but showed waning across all populations. Antibodies showed stable and equivalent levels of functional inhibition against spike-ACE2 binding in all age groups with comparable activity against viral variants of concern. SARS-CoV-2 seropositive donors showed high levels of antibodies to other beta-coronaviruses but serostatus did not impact humoral immunity to influenza or RSV. SARS-CoV-2-specific cellular responses were equivalent across the life course but virus-specific populations showed elevated levels of activation in older donors. LTCF residents who are survivors of SARS-CoV-2 infection thus show robust and stable immunity which does not impact responses to other seasonal viruses. These findings augur well for relative protection of LTCF residents to re-infection. Furthermore, they underlie the potent influence of previous infection on the immune response to Covid-19 vaccine which may prove to be an important determinant of future vaccine strategy.

**One sentence summery:** Care home residents show waning of nucleocapsid specific antibodies and enhanced expression of activation markers on SARS-CoV-2 specific cells

## Introduction

A striking feature of the current SARS-CoV-2 pandemic has been the high rates of mortality in older people. The biological basis for this observation is currently unclear but may relate to relative impairment of innate immune or adaptive responses and increased prevalence of co-morbidities. Long term care facilities (LTCF) provide residential and/or nursing care support for some of the frailest and elderly (>65) people within the population and as such have proven particularly vulnerable to the impact of SARS-CoV-2 infection(1). Rates of SARS-CoV-2 infection within LTCF have varied considerably and a range of factors have been determined that may impact susceptibility to infection(2). Mortality rates amongst elderly residents have been estimated at up to 30%(3) with factors such as cognitive impairment and extreme ageing as significant risk factors(3-5). Nevertheless, the majority of older people do recover from acute SARS-CoV-2 infection and this will be dependent on the generation of a functional SARS-COV-2-specific immune response. This will also help to provide protection against subsequent reinfection and will enhance immune responses at the time of Covid-19 vaccination. However, studies of staff and residents within LTCF are difficult to perform and at the current time there is very limited information regarding the nature of the SARS-CoV-2-specific response after natural infection in this setting. This is particularly important as these features may show differential features across the life course that could impact relative susceptibility to reinfection.

Vaccination against Covid-19 has proven highly effective and evidence to date shows that this protective effect is also observed within elderly and frail individuals(6, 7). As such, LTCF vaccination programmes offer the potential to limit the impact of the pandemic in this setting. However, here it will also be important to determine how the features of natural immunity impact the immune response to vaccination as SARS-CoV-2 serostatus impacts dramatically on the response to both single and double vaccination. These studies are required to determine the relative need for booster vaccination, particularly within vulnerable populations where there has been concern about the longevity of vaccine-induced immunity.

A further issue of importance relates to the potential impact of SARS-CoV-2 serostatus on heterologous immune responses to other pathogens. Seasonal respiratory viruses such as influenza and respiratory syncytial virus (RSV) remain a cause of considerable morbidity and mortality in older people and it is currently unclear if primary SARS-CoV-2 infection, with its associated acute inflammatory response, might act to suppress memory antibody responses against other pathogens, as has been seen in other settings. If so, this could have a considerable impact on policy decisions in relation to the annual influenza vaccination strategy. Furthermore, there is increasing concern about the long-term complication of ‘long-covid’ which may partly reflect a sustained systemic inflammatory profile after SARS-CoV-2 infection. This may be of particular importance in older people as many inflammatory markers increase naturally with age in a process that has been termed ‘inflamm-ageing’ and is particularly marked in those with chronic health conditions (8). To date, there is no information on how SARS-CoV-2 serostatus impacts systemic inflammatory markers in older people in the care home setting.

We determined virus-specific and general inflammatory profiles in both staff and residents in the LTCF setting over a 4-month period and related these findings to SARS-CoV-2 serostatus. Overall, robust SARS-CoV-2-specific and heterologous immune responses were observed across the life course which is encouraging for longer-term health outcomes and responsiveness to Covid-19 vaccination.

## Results

### SARS-CoV-2-specific antibody responses are higher in residents within LTCFs

Blood samples were collected from 276 staff and residents at England Long Term Care Facilities (LTCF) between June and November 2020, prior to the introduction of Covid-19 vaccination on 8th of December 2020 (Table 1). Baseline samples were collected between June and July with matched follow up samples at 2 and 4 months later. 163 donors were found to be SARS-CoV-2 seropositive based on N-antibody status and 113 were seronegative. The peak time point for primary SARS-CoV-2 infection within LTCF was April 2020 and as such these time points likely represent time points up to 7 months after primary infection.

**Table 1.**
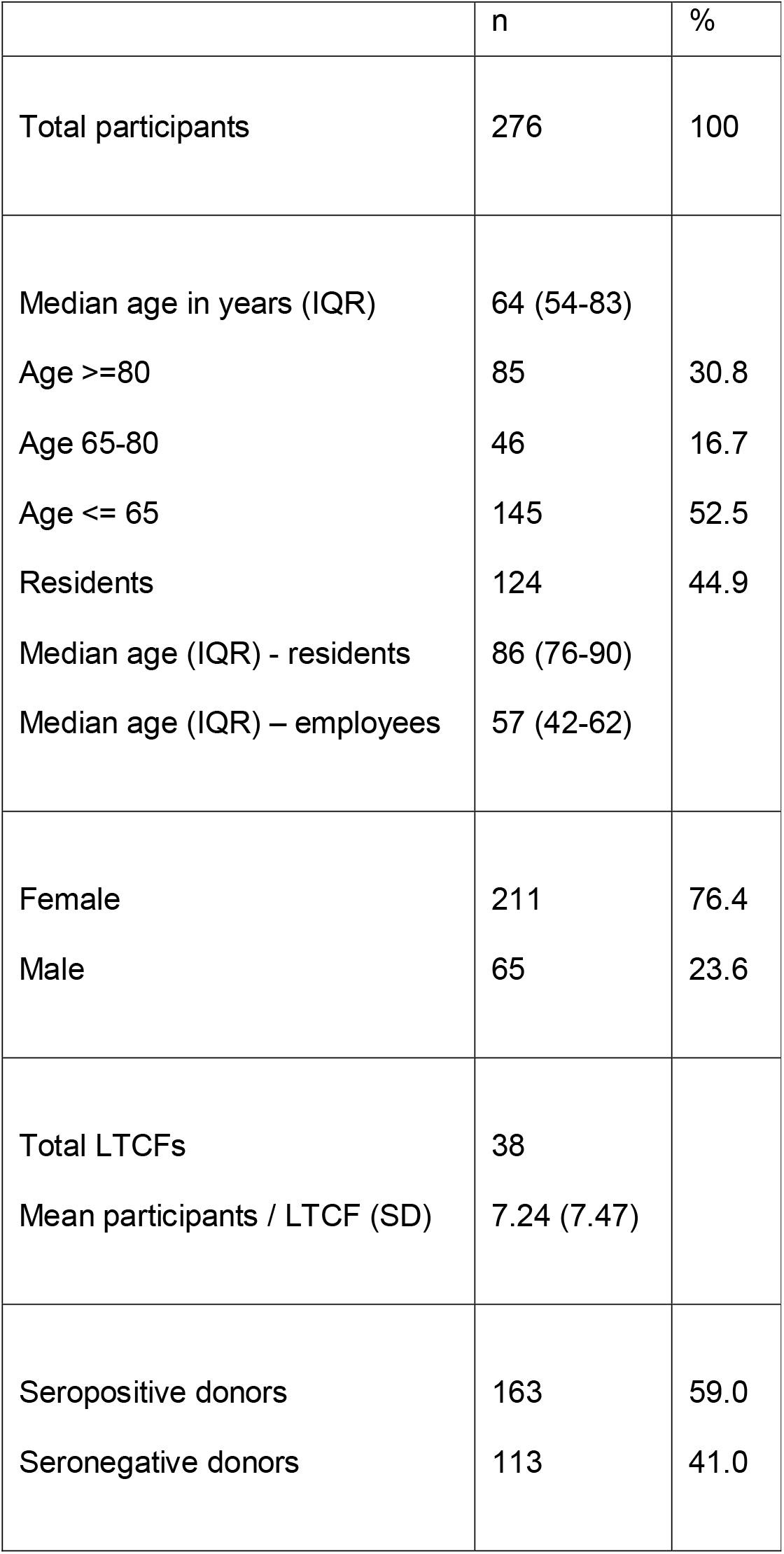
Demographics of donors.

|  | n | % |
| --- | --- | --- |
| Total participants | 276 | 100 |
| Median age in years (IQR) | 64 (54-83) |  |
| Age >=80 | 85 | 30.8 |
| Age 65-80 | 46 | 16.7 |
| Age <= 65 | 145 | 52.5 |
| Residents | 124 | 44.9 |
| Median age (IQR) - residents | 86 (76-90) |  |
| Median age (IQR) – employees | 57 (42-62) |  |
| Female | 211 | 76.4 |
| Male | 65 | 23.6 |
| Total LTCFs | 38 |  |
| Mean participants / LTCF (SD) | 7.24 (7.47) |  |
| Seropositive donors | 163 | 59.0 |
| Seronegative donors | 113 | 41.0 |

Initial studies focused on assessment of SARS-CoV-2-specific humoral immunity in the 163 seropositive staff and residents. MSD technology was used to determine the magnitude of spike-specific, RBD-specific and nucleocapsid (N)-specific antibody responses and data were compared between four age quintiles: <40; 40-64; 65-85 and 85+ years of age (n=19, 67, 42 and 35 respectively). Serial analysis at 2 and 4 months after the initial collection allowed assessment of antibody waning during this period.

Spike-specific and RBD-specific antibody responses were both higher in LTCF residents aged 65+ years compared to younger donors with values that were 1.7-times and 2.1-times increased respectively (p=0.004 and p=<0.0001 respectively) (Fig 1a-d). Nucleocapsid-specific responses were also increased in the elderly donors aged 85+ years where they were 2.2-times higher than donors aged <65 years (p=0.04) (Fig 1e/f).

**Figure 1:**
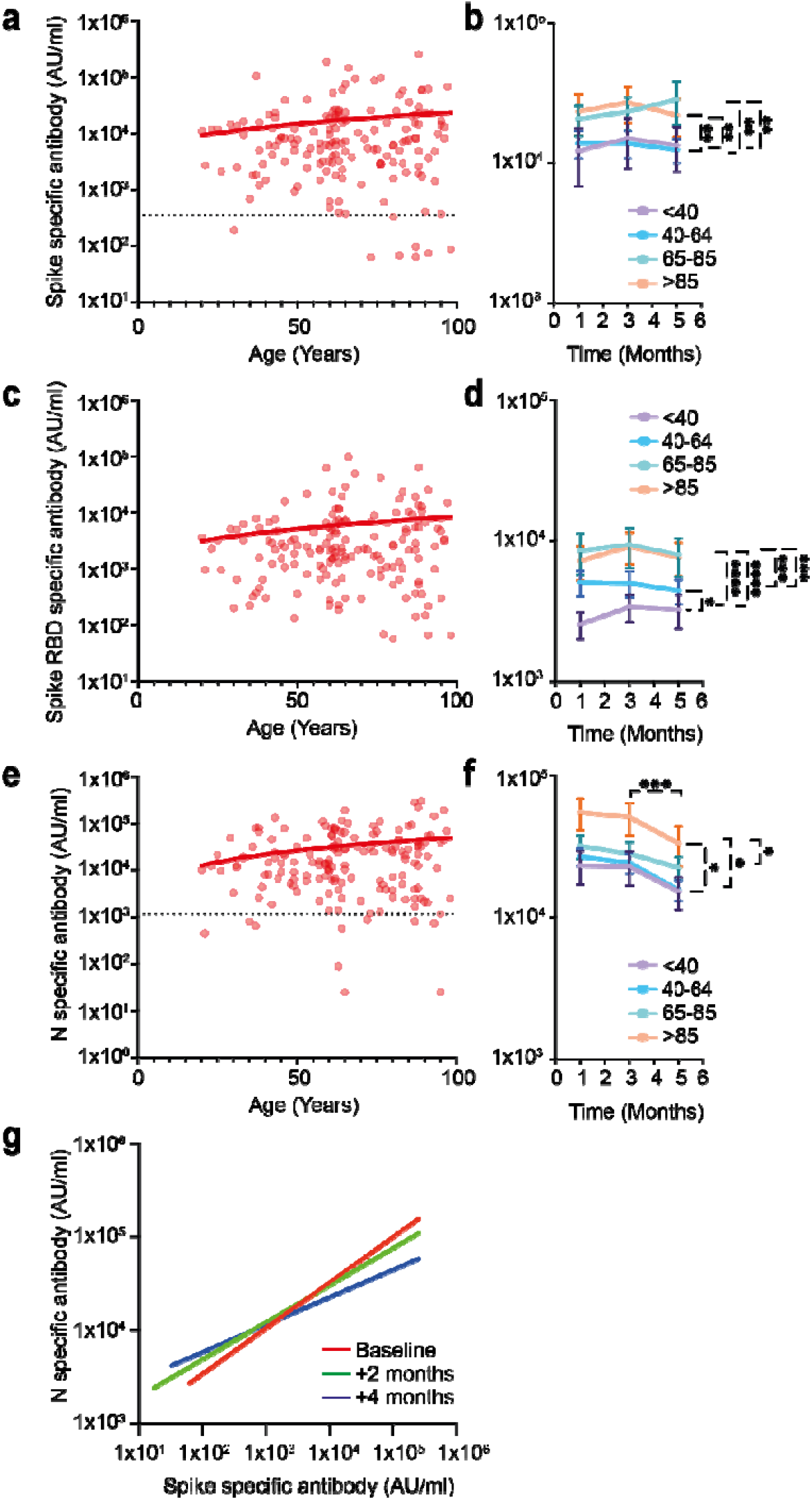
Elevated levels of SARS-CoV-2 Spike and Nucleocapsid-specific antibodies in LTCF residents. a) SARS-CoV-2 Spike-specific IgG response in relation to age within seropositive donors (n=163) (r=0.11, p=0.16). Dotted black line indicates cut-off. b) Mean spike-specific antibody responses at baseline and 2 and 4 month follow up within 4 age groups, <40 (purple), 40-64 (blue), 65-85, (green) and >85 years (orange). Error bars indicate standard error. ^**^= p <0.001. c) SARS-CoV-2 RBD-specific IgG response in relation to age in seropositive donors (n=163) (r=0.11, p=0.17). d) Mean RBD-specific antibody responses at baseline and 2 and 4 month follow up within 4 age groups, <40 (purple), 40-64 (blue), 65-85, (green) and >85 years (orange). Error bars indicate standard error. ^*^= p<0.05, ^**^= p 0.002, ^***^= p=0.0006, ^****^=p<0.0001. e) SARS-CoV-2 Nucleocapsid-specific IgG response in relation to age within seropositive donors (n=163) (r=0.19, p=0.01). Dotted black line indicates cut-off. f) Mean nucleocapsid-specific antibody responses at baseline and 2 and 4 month follow up within 4 age groups, <40 (purple), 40-64 (blue), 65-85, (green) and >85 years (orange).. Error bars indicate standard error. ^*^= p<0.05, ^***^= p=<0.01. g) Correlation between SARS-CoV-2 Spike and Nucleocapsid-specific antibody responses in all donors at 3 time points, baseline (red) (r= 0.26), 2 months after baseline (green) (r= 0.17) and 4 months after baseline (blue) (r=0.08) (n=115). The lines of best fit are shown.

The potential importance of antibody waning was next assessed in relation to donor age. Temporal assessment of antibody responses over four months showed that antibody responses against SARS-CoV-2 spike and RBD were stable over time (Fig 1g). This is encouraging in relation to potential protection against reinfection and supports clinical assessment in relation to the capacity of natural immunity to prevent infection.

In contrast, relative waning of antibody responses against nucleocapsid protein was observed in all age groups during the 4 months of study. This was particularly marked in the older donors where mean titres fell by 39% during the 4-months of follow up (p=0.01) (Fig 1e-g). This pattern of enhanced antibody waning against nucleocapsid has been seen in other settings (9) and was reflected in the spike: nucleocapsid antibody ratio in donors aged >85+ years which decreased from 2.4-times at baseline to 1.6 times at 4 months of follow up (p=0.0003) (Fig 1f and supplementary table 1).

### SARS-CoV-2-specific antibody responses demonstrate comparable and stable functional profile to SARS-CoV-2 variants of concern in staff and residents

In order to assess the functional capacity of antibodies elicited after natural infection across the life course we next investigated the capacity of antibodies from both staff and residents to inhibit the binding of soluble recombinant spike protein to ACE2. Importantly, in addition to the prototypic Wuhan sequence, recombinant spike protein from the viral Variants of Concern (VOC) B.1.1.7 (alpha), B.351 (beta) and P.1 (gamma) was also incorporated in the assay.

Immune sera were markedly impaired inability to inhibit spike-ACE2 binding for VOC compared to the parental Wuhan virus (34% SD 21.7%). This was particularly true for the B.351 (16.6%, p=<0.0001) and P.1 (15.4%, p=<0.0001) VOC whilst inhibition of B.1.1.7 variant binding (29.4%, p=0.78) was comparable to Wuhan in both assays (Fig 2a). Of note, we also observed that this profile of VOC spike inhibition was stable over 4 months of follow up which indicates that the functional activity of antibodies does not decline (Fig 2a). Furthermore, no differences were observed in relation to the pattern of spike-inhibition using serum samples from donors across all ages (data not shown).

**Figure 2:**
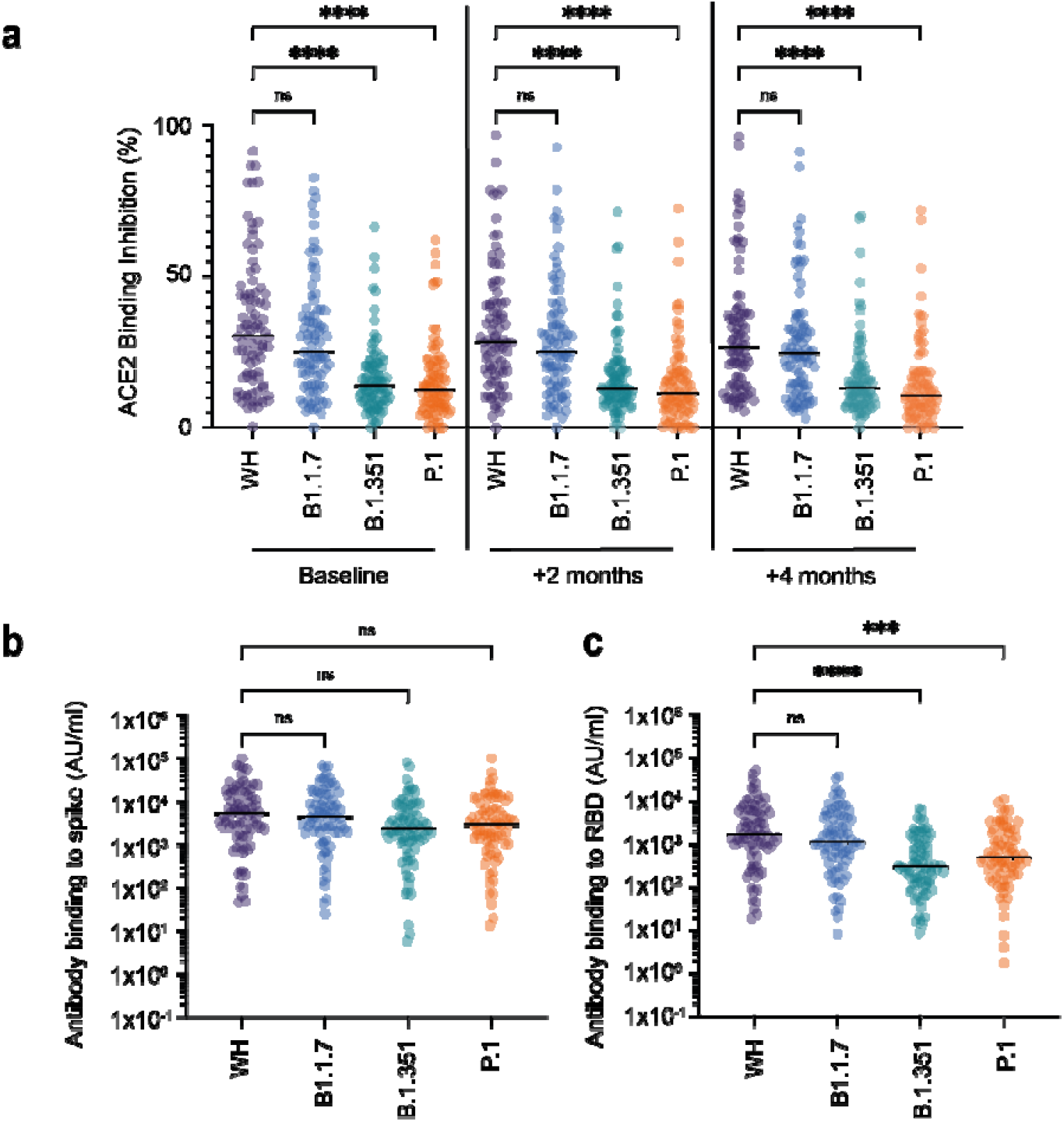
Serological inhibition of spike-ACE2 interaction and direct antibody binding to Spike/RBD. a) Serological inhibition of spike binding to ACE2. Spike proteins were available from viral variants Wuhan (WH) (purple), B1.1.7 (blue), B.1.351 (green) and P.1 (orange). Sera were taken from donors after natural infection at baseline, 2 and 4 months after baseline sample. ^****^=p<0.0001 (n=163). b) Serological binding to SARS-CoV-2 spike from viral variants (Wuhan [WH] [purple], B1.1.7 [blue], B.1.351 [green] and P.1 [orange]) after natural infection at baseline. N.S = not significant (n=163). c) Serological binding to SARS-CoV-2 RBD from viral variants (Wuhan [WH] [purple], B1.1.7 [blue], B1.351 [green] and P.1 [orange]) after natural infection at baseline. ^****^ = p<0.0001 (n=163).

We next determined direct serum binding to spike proteins from VOC. Of interest, antibody binding to total spike protein was comparable against all four VOC indicating that the mutational changes within each VOC do not significantly change the total antibody-binding capacity of the spike protein (Fig 2b). However, serological neutralisation of virus binding to ACE2 is mediated primarily through binding to RBD and here binding was substantially reduced against both B.351 (842 AU/ml, p=<0.0001) and P.1 (1377 AU/ml, p=0.0003) compared to 5170 AU/ml against the Wuhan prototype (Fig 2c). This profile was in line with the pattern of Spike-ACE2 inhibition and again this pattern was both stable over time and across the age groups (data not shown).

These data show that the functional capacity of spike-specific antibodies elicited after natural infection is stable over at least four months and is independent of age in the LTCF setting.

### Prior history of SARS-CoV-2 infection leads to boosting of antibody responses against beta coronaviruses

We next went on to assess how previous infection with SARS-CoV-2 impacted on the magnitude of the antibody response against other human coronaviruses. The MSD platform was used to assess binding to spike protein from the beta-coronaviruses SARS-CoV-1, HKU1 and OC43 as well as the alpha-coronaviruses 229E and NL63.

As anticipated, antibody binding to SARS-CoV-1 was very low in SARS-CoV-2-seronegative individuals but substantially increased within seropositive donors (10) due to substantial sequence homology between viruses. This was strongly enhanced in older donors reflecting the elevated SARS-CoV-2-specific response in this age group (r=0.16 p=0.03) (Fig 3A).

**Figure 3:**
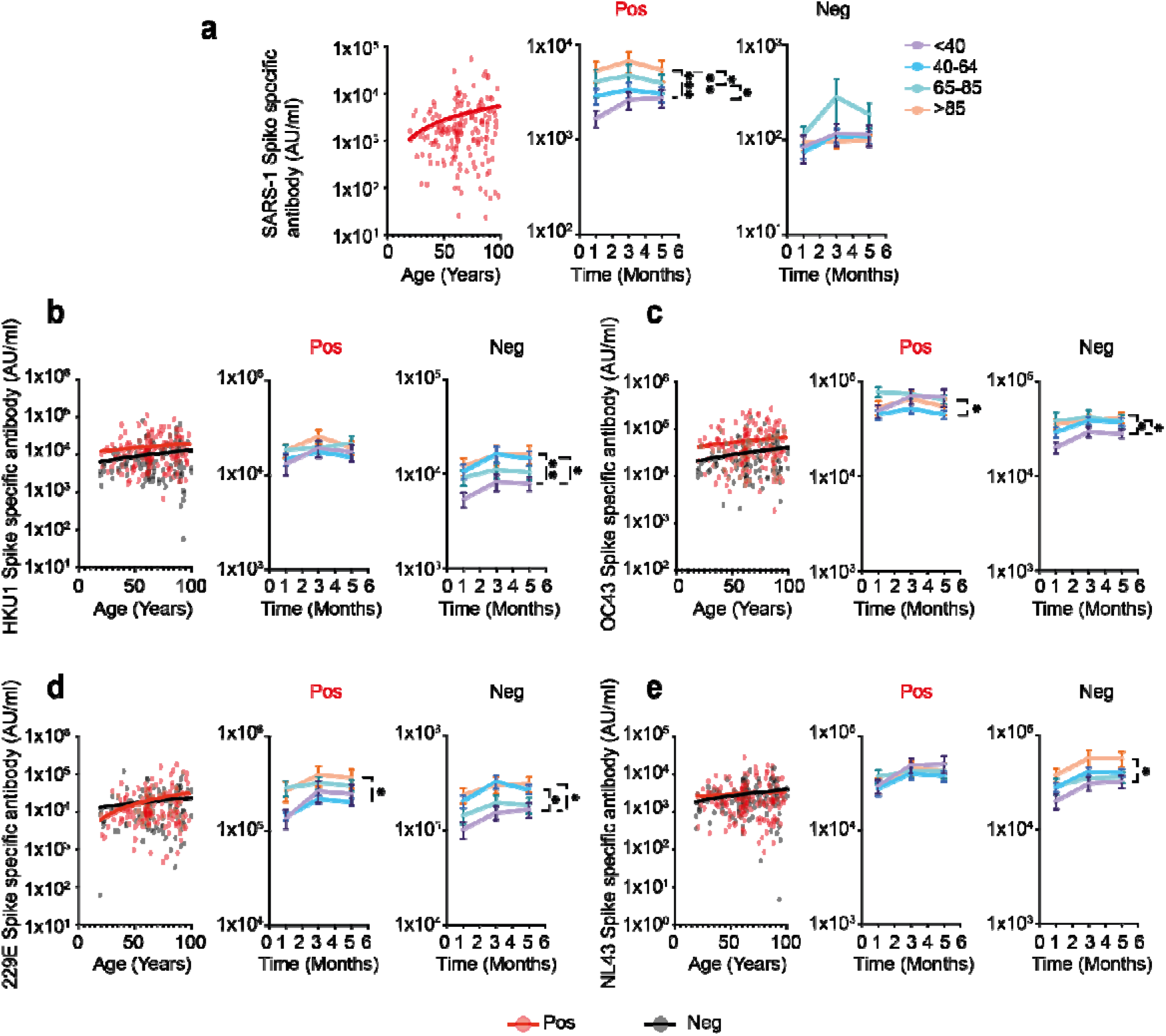
IgG antibody responses against beta-coronaviruses are increased in donors with previous SARS-CoV-2 infection. Comparison of antibody responses to coronaviruses in relation to age and SARS-CoV-2 serostatus. Seronegative donors are indicated by black dots (n=113) and seropositive donors (n=163) are indicated by red dots. Lines indicate line of best fit. Stability of responses over 4 months of follow up is shown within four age ranges; <40 (purple), 40-64 (blue), 65-85, (green) and >85 years (orange). Error bars indicate standard error. a) SARS-1 Spike-specific antibody response in SARS-CoV-2 seropositive donors (r=0.16 p=0.03). Stability of response in age cohorts is shown on the right. *p=0.035 ^**^p =0.0013 ^***^p=0.0003. b) HKU-1 Spike-specific antibody response. (Seropositive: r= 0.10, p=0.16; Seronegative: r=0.14, p =0.13). Stability of response in age cohorts is shown on the right: ^*^p=0.01 ^**^p= 0.008. c) OC43 Spike-specific antibody response. (Seropositive: r= 0.11, p=0.15; Seronegative: r=0.17, p=0.06). Stability of response in age cohorts is shown on the right: seropositive ^*^ p=0.01; seronegative ^*^p=<0.02. d) 229E Spike-specific antibody response (Seropositive: r=0.23, p=0.003; Seronegative: r=0.13, p =0.16). Stability of response in age cohorts is shown on the right: seropositive p=0.02; seronegative ^*^p=<0.03. e) NL63 Spike-specific antibody response (Seropositive: r= 0.06, p=0.42; Seronegative r=0.19, p =0.03). Stability of response in age cohorts is shown on the right: seronegative ^*^p=<0.02.

However, it was also noteworthy that enhanced antibody binding was observed against both of the endemic beta-coronaviruses in SARS-CoV-2 positive donors (OC43 55451 AU/ml vs 31539 AU/ml, p=<0.0001 and HKU1 16223 AU/ml vs 10083 AU/ml, p=0.0008) (Fig 3b/c). Of note, OC43-specific responses were also enhanced in seronegative older people (r=0.17 p =0.06), potentially reflecting the impact of recurrent priming from seasonal infection (Fig 3b/c).

Alpha-coronaviruses show less structural homology with SARS-CoV-2 spike protein and this may underlie the finding of no increment in serological binding to spike protein in SARS-CoV-2 seropositive donors. However, the serological response to alpha-CoV was again increased in older people prior to SARS-CoV-2 infection (NL63: r=0.19 p =0.03) but this differential was lost following seroconversion (Fig 3d/e).

These findings show that serological responses against both alpha and beta-coronaviruses increase with age, potentially due to recurrent infections. However, cross-reactive antibody responses are increased following SARS-CoV-2 infection and this effect is both stronger in younger people and more marked for responses against the beta-coronaviruses. The relationship of these observations to clinical protection against individual coronaviruses is uncertain at present.

### SARS-CoV-2 serostatus does not impair antibody responses against other respiratory viruses

Acute respiratory infections such as influenza and respiratory syncytial virus (RSV) represent a major and continuous challenge to the health of elderly individuals and annual vaccination against influenza is recommended in many countries. Protection against infection is believed to be mediated primarily through the action of humoral immunity and there is concern that SARS-CoV-2 serostatus may impact on the titre of heterologous immune responses. As such, we next assessed the strength of antibody responses against a range of influenza viral strains in relation to age and SARS-CoV-2 serostatus.

SARS-CoV-2 serostatus was not seen to have any influence on the titre of influenza or RSV-specific antibody responses across the different age groups (Fig 4). Relative antibody titres against influenza were broadly stable across the life course whilst the serological response against RSV increased with age.

**Figure 4:**
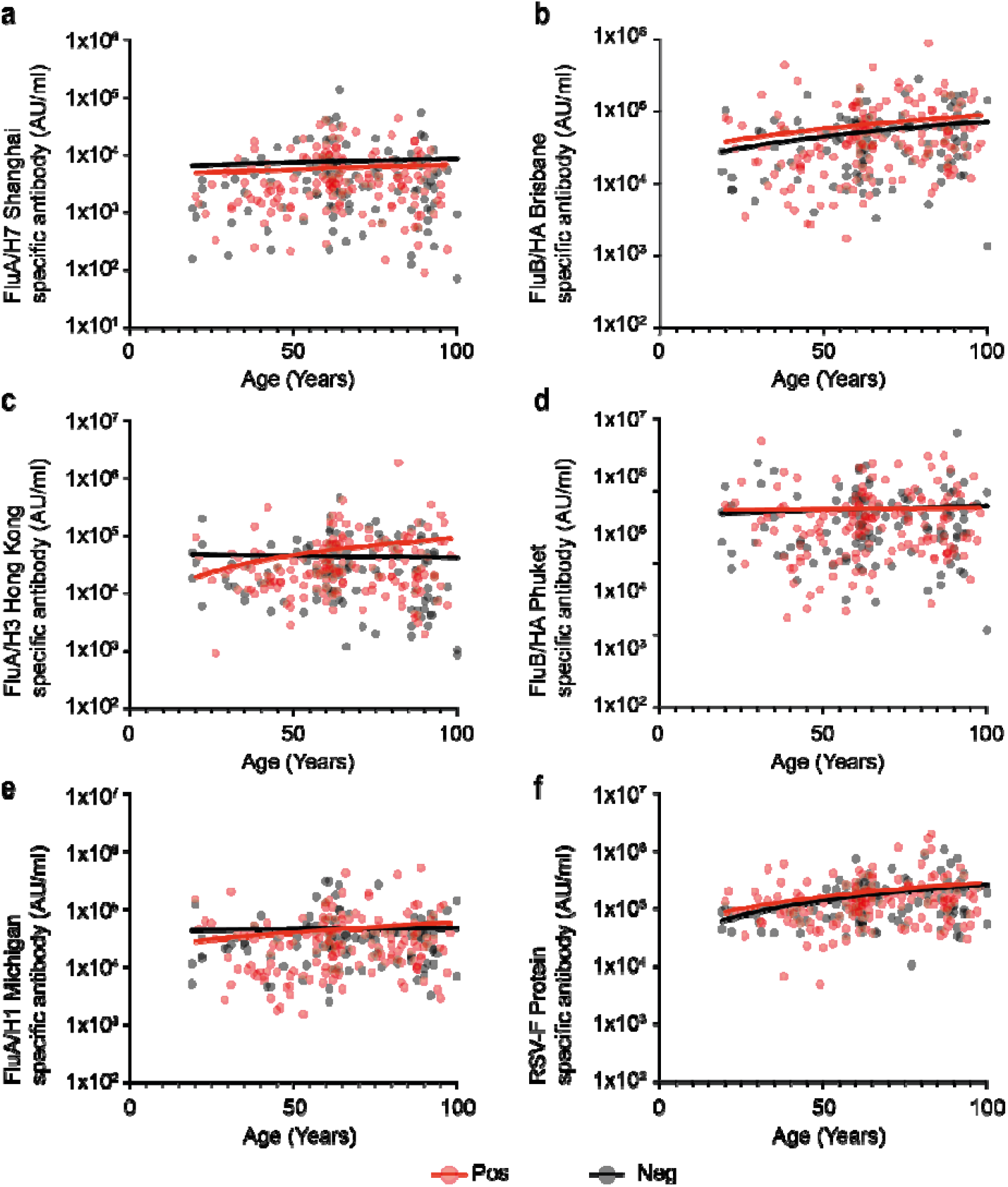
SARS-CoV-2 serostatus does not influence IgG responses to other respiratory viruses. Comparison of IgG response against respiratory viruses in relation to age and SARS-CoV-2 serostatus. Red dots indicate positive donors (n=164) and black dots indicate negative donors (n=113). Statistical analysis below is shown for seropositive and seronegative donors respectively. a) Influenza A/H7 Shanghai (r=0.06, p=0.40; r=0.03 p=0.72). b) Influenza B/HA Brisbane (r=0.12, p=0.10; r=0.22 p=0.15). c) Influenza A/H3 Hong (r=0.11, p=0.16; r=-0.02 p=0.77). d) Influenza B/HA Phuket (r=0.18, p=0.81; r=-0.04 p=0.64). e) Influenza A/H1 Michigan (r=0.10, p=0.18; r=-0.01 p=0.85). f) RSV-F protein (r=0.18, p=0.02; r=-0.29 p=0.001).

These data provide reassurance that previous history of SARS-CoV-2 infection does not impact detrimentally on the profile of immunity against heterologous respiratory viruses such as influenza or RSV.

### SARS-CoV-2-specific cellular responses are comparable between staff and residents

Cellular immune responses are likely to play an important role in protection against reinfection with SARS-CoV-2. As such, IFN-γ ELISpot analysis was next used to determine T cell responses against both a peptide pool from spike protein and a combination of peptides from nucleocapsid, membrane and envelope protein (N/M/E). Analysis was performed in 80 donors and assessed in three age groups of <40, 40-64 and 65+ years.

The magnitude of the ELISpot response against spike peptides was comparable in donors at all ages and was also stable over the four months of follow up (Fig 5a-c). In contrast, cellular responses against the N/M/E pool were lower in donors aged under 40 years (36 SFU/10^6^) when compared to 40-64s (129 SFU/10^6^ p=0.02) and >65+ age groups (167 SFU/10^6^ p=0.005) (Fig 5d-f).

**Figure 5:**
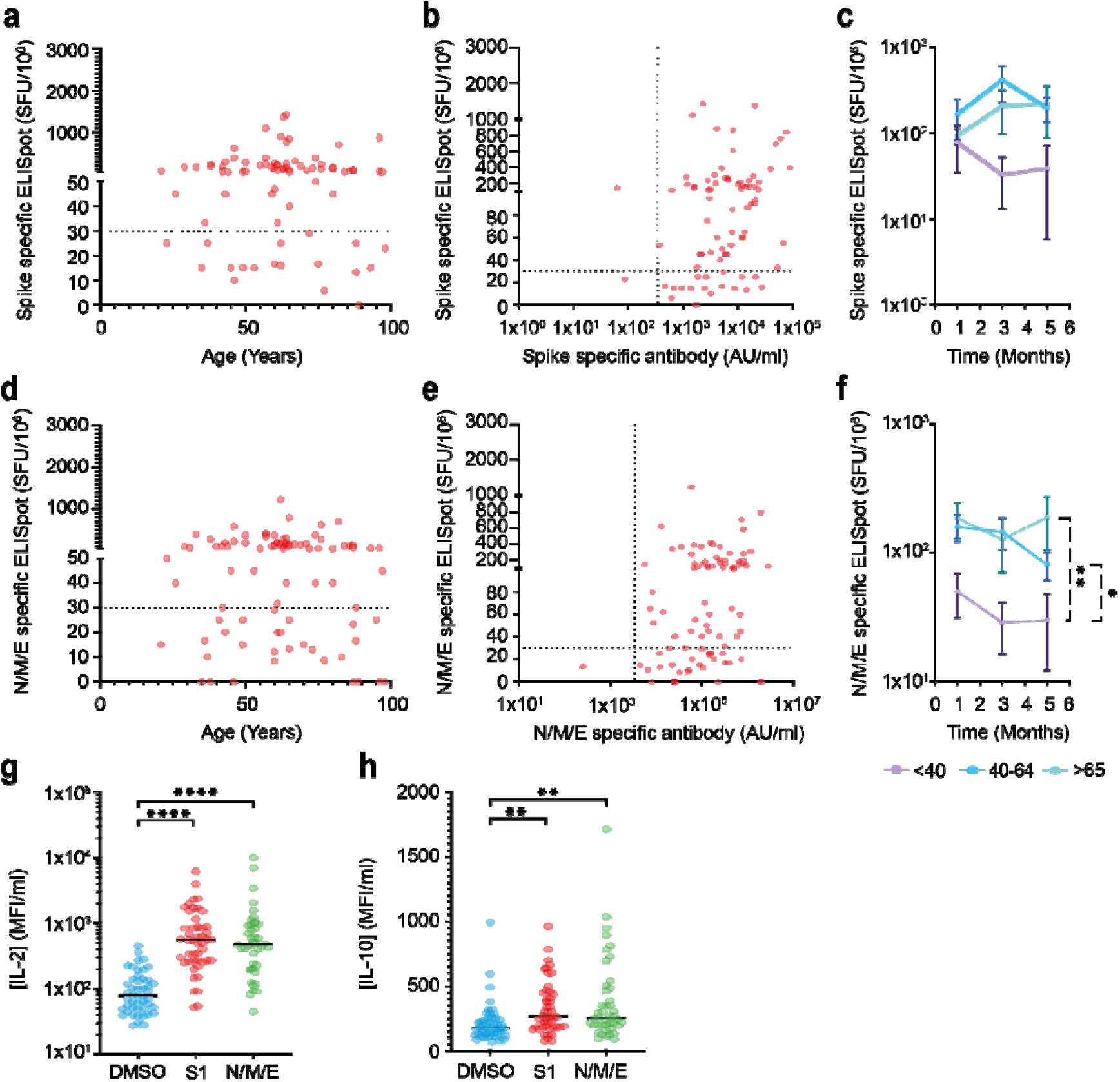
SARS-CoV-2-specific T cell responses are observed across the lifecourse and demonstrate IFN-gamma, IL-2 and IL-10 production. a) Spike-specific cellular response in relation to age in seropositive donors (n=80) (r= 0.02 p= 0.82). Dotted black line indicates cut off for positive response. b) Comparison of spike-specific cellular and antibody responses in seropositive donors (n=80) (r=0.32 p=0.003). c) Mean spike-specific cellular responses within age categories <40 (purple), 40-64 (blue), and >65 years (green) at 0, 2 and 4 months. Error bars indicate standard error. d) N/M/E-specific cellular response in relation to age in seropositive donors (n=80) (r= -0.03 p= 0.72). Dotted black line indicates cut off for positive response. e) Comparison of N/M/E-specific cellular and antibody responses in seropositive donors (n=80) (r=0.35 p=0.001). f) Mean N/M/E-specific cellular responses within age categories, <40 (purple), 40-64 (blue), and >65 years (green) at 0, 2 and 4 months. Error bars indicate standard error. ^*^ p=0.02 ^**^p=0.005 g) IL-2 concentration in ELIspot supernatants in DMSO (blue), S1 (red) and N/M/E (green) peptide stimulated wells (n=48). ^****^p=<0.0001 h) IL-10 concentration in ELIspot supernatants in DMSO (blue) S1 (red) and N/M/E (green) peptide simulated wells (n=48). ^**^p=<0.002

To further assess the functional profile of SARS-CoV-2-specific cells we then measured the concentration of cytokines within ELIspot eluates using LEGENDplex technology. Markedly increased concentrations of IL-2 and IL10 were seen in both the spike and N/M/E-specific eluates compared to DMSO control (Fig 5g-h) (Spike: IL-2; 110 MFI/ml (control) vs 890 MFI/ml, p=<0.0001; IL-10: 220 MFI/ml (control) vs 330 MFI/ml, p=0.002); (N/M/E: IL2; 110 MFI/ml (control) vs 1000 MFI/ml, p=<0.0001; IL-10: 215 MFI/ml (control) vs 380 MFI/ml, p=0.0019).

### SARS-CoV-2-specific CD4+ T cells are present at low frequency and show increasing expression of HLA-DR with age

Covid-19 elicits a robust T cell response, but it remains unclear how the phenotype of this response varies across the life course or how this may modulate the overall T cell repertoire. This is of particular note given the accumulation of effector T cells and enhanced inflammatory profile associated with ageing. As such, we next used 20-plex fluorometric analysis to determine expression of a range of memory, differentiation and activation markers across the global T cell repertoire both at baseline and at 4 months of follow up (n=65; supplementary table 2). tSNE analysis revealed a broadly comparable profile within the peripheral T and NK cell repertoire both at baseline and follow up (Figure 6a).

**Figure 6:**
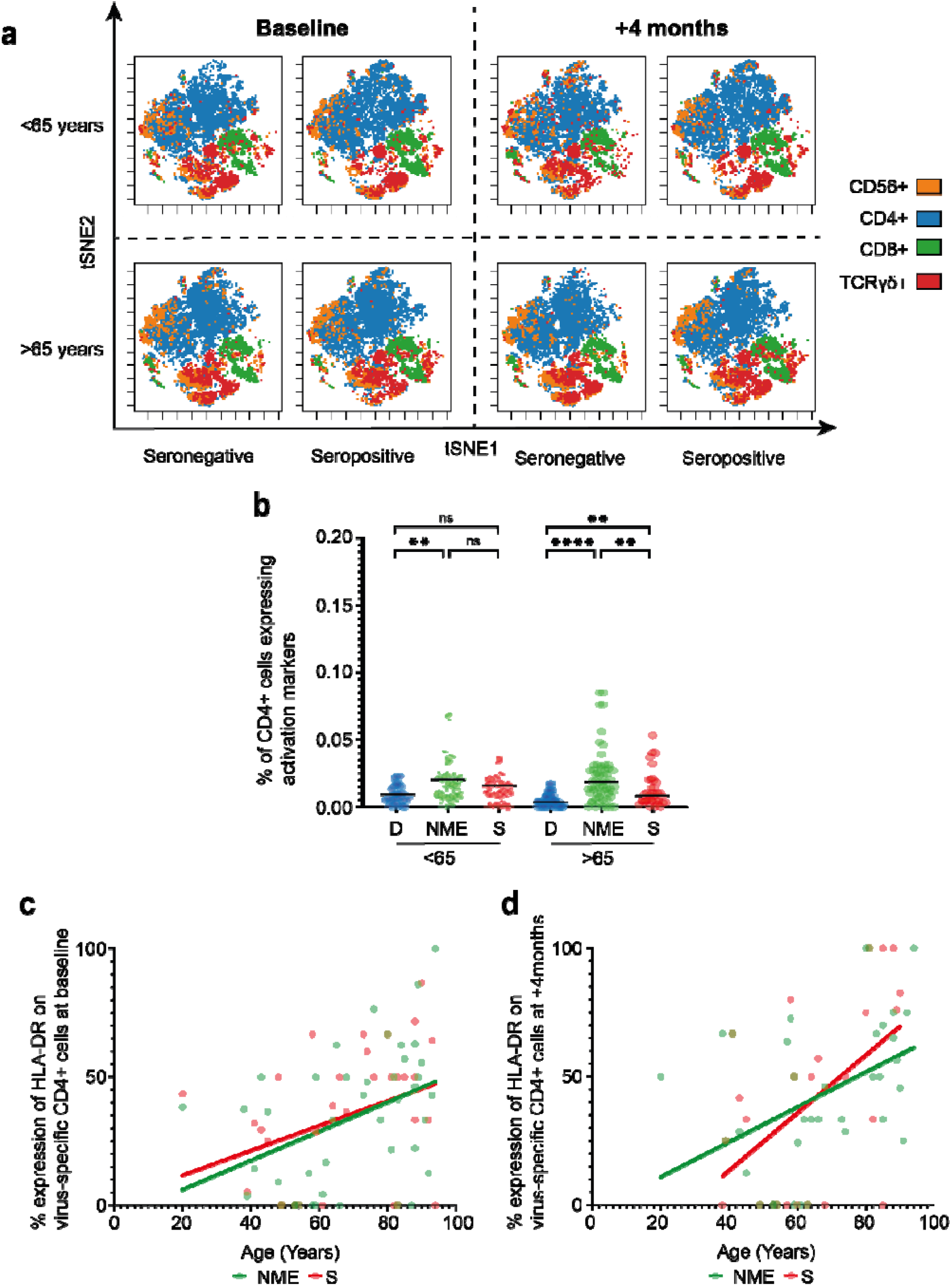
Global and SARS-CoV-2-specific T cell response within staff and residents in LTCF. a) t-SNE plots of global T and NK cell repertoire in younger and older donors in relation to SARS-CoV-2 serostatus. Expression of CD4+, CD8,C D56+ and TCRγδ+ is shown to demonstrate major cell lineages. CD19 and CD14 cells were excluded from analysis. ∼20,000 PBMCs were identified using integrated and classification analysis and data from both baseline samples and +4 month samples is shown. b) Quantification of SARS-CoV-2-specific T cells based on percentage expression of both CD137+ and CD154+ on CD4+ cells after stimulation with N/M/E or spike (S) peptides. DMSO (D) is used as a negative control. Analysis is grouped into staff (<65) or residents (>65 years). Black line indicates the median. ^**^ p= >0.002 ^****^ p= <0.0001. c,d) Percentage expression of HLA-DR+ on CD137+CD154+CD4+ cells following stimulation with N/M/E (r=0.40 p=0.006; n=44) or spike peptides (r=0.36 p=0.03; n=33) in relation to age at baseline (c) and at 4 months (d).

Activation-induced marker (AIM) assessment was used to determine the magnitude and profile of the SARS-CoV-2-specific immune response following stimulation with viral peptide pools (n=44) (supplementary figure 1). The magnitude of SARS-CoV-2-specific T cell response was modest and comparable across age groups (fig 6b). In particular, within LTCF staff a median of 0.02% and 0.016% of CD4+ cells demonstrated specificity for N/M/E or spike respectively. Comparable values within the resident population were 0.018% and 0.008% respectively. (fig 6b), comparable with previous reports. Virus-specific CD8+ cells were not detected with this approach(11).

Expression of memory and differentiation markers on the virus-specific pool demonstrated a dominant CD27+CD28+CD127^low^ CD4+ effector phenotype (data not shown). Of note, expression of the activation marker HLA-DR increased substantially with age on both spike and N/M/E-specific CD4+ T cells (spike: r=0.36 p=0.03, N/M/E: r=0.40 p=0.006) (fig 6c/d). This profile was present at baseline and was further accentuated on the spike-specific pool at the four-month time point where a median of 75% of cells within residents expressed HLA-DR compared to 25% of cells with staff (n=28; p=0.02)(Figc-d). CD95 (Fas) expression was also similar between but did increase during follow up (supplementary figure 2). Expression of CD25, an additional marker of activation, did not show any such relationship with age (data not shown)

These data indicate that SARS-CoV-2-specific T cells comprise a small component of the peripheral T cell pool but virus-specific cells in older people show increased expression of HLA-DR suggesting chronic activation.

### Serum inflammatory cytokine levels increase with age and are partially attenuated by SARS-CoV-2 infection

Acute primary infection with SARS-CoV-2 increases serum concentration of many inflammatory cytokines such as IL-6, TNF-α and IFN-γ. Sustained elevated levels of these proteins have been seen in a subgroup of patients with more prolonged disease and as such we next went on to assess systemic cytokine concentration in relation to SARS-CoV-2 serostatus across the life course.

MSD analysis was used to measure concentrations of IFN-γ, TNF-α, IL-6, IL-2 and IL-10 in plasma samples (Fig 7). Concentrations of the inflammatory cytokines IL-6, IFN-γ, IL-2 and TNF-α increased with age within SARS-CoV-2 seronegative donors and likely reflect the established profile of ‘inflammageing’(8). Of interest, within SARS-CoV-2 seropositive donors this association was lost for IFN-γ and IL-2, suggesting that previous SARS-CoV-2 infection may moderately suppress plasma concentrations in older people.

**Figure 7:**
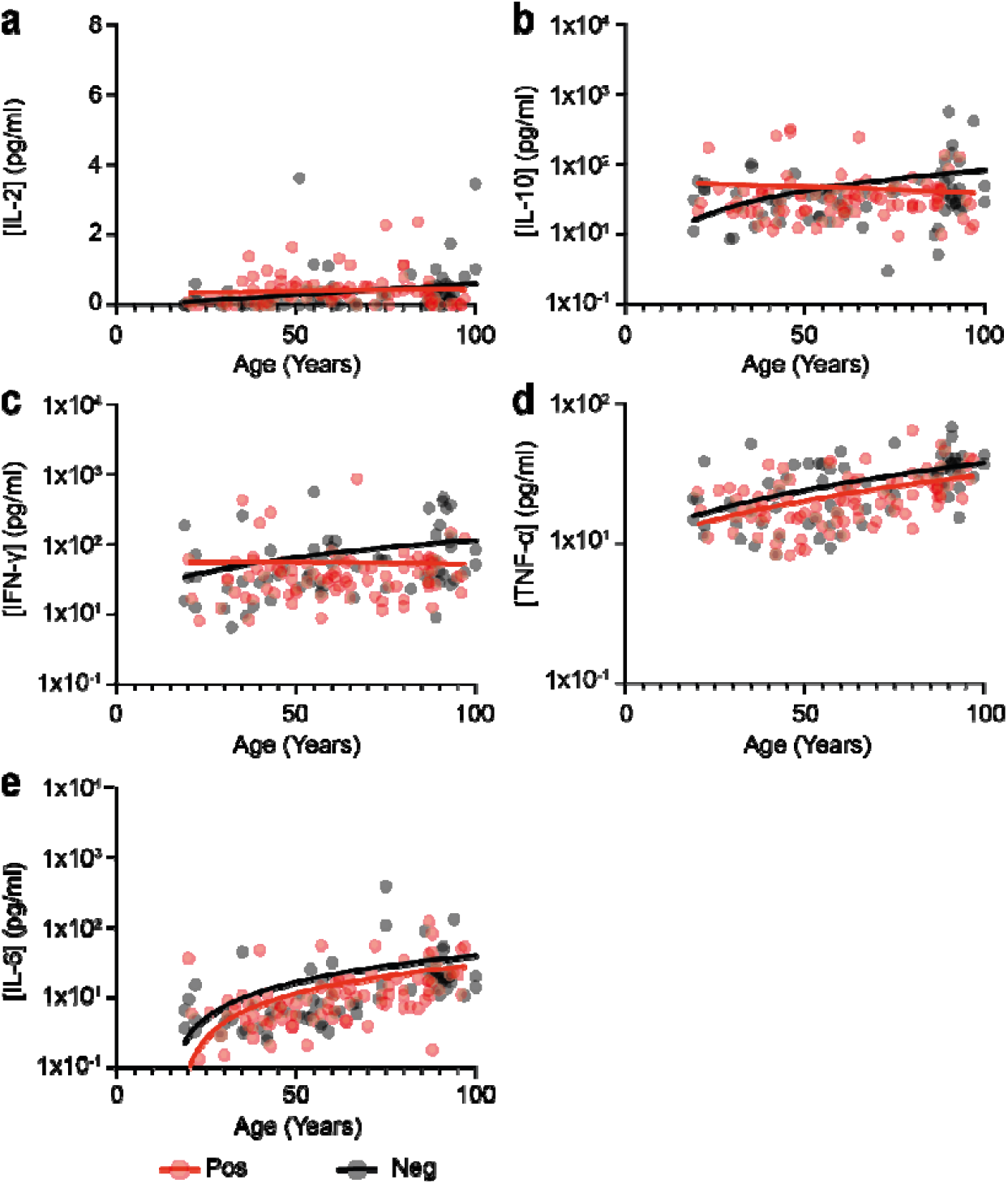
SARS-CoV-2 serostatus may partially attenuate inflammageing. Comparison of plasma cytokine concentration in relation to age and SARS-CoV-2 serostatus. Black dots indicate seronegative donors (n=72) and red dots indicate seropositive donors (n=93). Statistical analysis assesses concentration in relation to age and is shown for seronegative and seropositive donors respectively. a) IL-2 (r=0.41 p=0.0003; r=0.008; p=0.93). b) IL-10 (r=0.20 p=0.08; r=0.11; p=0.28). c) IFN-γ (r=0.46 p=<0.0001; r=0.18; p=0.07). d) TNF-α (r=0.52 p=<0.0001; r=0.47; p=<0.0001). e) IL-6 (r=0.67 p=<0.0001; r=0.59; p=<0.0001).

These findings indicate that previous infection with SARS-CoV-2 does not lead to prolonged systemic inflammation and may even act to attenuate these levels in the longer term.

## Discussion

The SARS-CoV-2 pandemic has presented a major challenge to the management of Long Term Care Facilities with high rates of mortality amongst older and frail residents. Little is known regarding the profile of SARS-CoV-2-specific immunity or the potential impact of previous infection on heterologous immunity. Here we investigated SARS-CoV-2-specific immune responses in staff and residents of LTCF and found robust responses against Spike protein across all age groups and no negative impact on immunity to other respiratory viruses. These findings are broadly reassuring for future management decisions.

Antibody responses against the SARS-CoV-2 spike protein and RBD domain are thought to be critical in prevention against reinfection. As such we were encouraged to find that antibody responses against spike and RBD were robust in older people. Indeed, antibody titres were increased in people over 65 years of age. Nucleocapsid-specific antibody responses were also higher in LTCF residents. The reasons for this are not clear but may potentially reflect an elevated residual antibody ‘setpoint’ after enhanced response to higher levels of viral load at the time of primary infection in older individuals. Nevertheless, these findings are in line with the 85% protection against re-infection that is seen within the resident population, a level of protection somewhat higher than the value of 60% within staff (12, 13)

Antibody responses against endogenous alpha and beta-human coronaviruses show marked waning within the first year after infection and this has led to concern that a similar process may occur following SARS-CoV-2 infection(14). Reassuringly, we observed that antibody responses against both spike and RBD were well maintained for at least four months. These data are in line with reports showing stability of spike-specific responses in younger donors(15). In contrast, antibody responses against nucleocapsid protein showed significant waning in all age groups and represent a potential future challenge for identification of individuals with prior natural infection. The physiological importance of the nucleocapsid-specific antibody response in relation to protection against reinfection is somewhat unclear. This profile of differential waning of nucleocapsid and spike-specific antibody responses has been observed in other cohorts(16) and may reflect alternative mechanisms of antigen presentation at the time of primary infection, with spike protein dominant on the viral envelope and nucleocapsid present in high numbers within the virion. Furthermore, retention of spike protein has been demonstrated within the gastrointestinal tract and may serve to drive maintenance and somatic hypermutation of spike-specific humoral responses (17).

In addition to the magnitude of the humoral response we were also interested to assess the relative functional activity of antibodies in relation to age. Again, this was comparable in both staff and residents with comparable inhibition of binding of Wuhan virus spike protein to the ACE2 receptor. Inhibition was also assessed against SARS-COV-2 viral Variants of Concern (VOC) where reduced activity was seen against both the B.1.351 and P.1 variants, in line with previous reports(18), but was equivalent in staff and residents. Furthermore, binding inhibition remained stable over 4 months, indicating maintenance of antibody avidity. It is worth mentioning that seropositive participants were infected in the first wave of the pandemic before any known VOCs, therefore it is most likely that they were infected with the Wuhan strain.

Infection with heterologous viruses such as influenza and RSV remains a major health concern for LTCF residents and there is currently no information on how SARS-CoV-2 serostatus may impact on memory responses against these respiratory viruses. This is of particular note as during the pandemic there has been a low incidence of non-COVID respiratory infections due to social distancing and there is concern that rapid re-emergence of influenza and RSV infections may emerge as LTCF start to reduce use of disease control measures. Infection with viruses such as measles can erode antibody responses against other pathogens (19, 20), potentially due to displacement of plasma cells within the bone marrow, and the strong inflammatory response induced by primary SARS-CoV-2 infection may pose a similar risk. Again, it was encouraging to see that the breadth and magnitude of the human response against influenza subtypes were entirely comparable across donors irrespective of SARS-CoV-2 infection status. This provides reassurance that residents who have undergone SARS-CoV-2 infection will not show increased vulnerability against seasonal respiratory infections. It will now be of interest to assess the relative response following annual influenza vaccination.

The importance of cellular immunity in relation to the protection against SARS-CoV-2 reinfection remains uncertain but it is likely that cellular responses play a critical role in protection against severe disease. Impairment of natural and vaccine-induced cellular immune responses is a feature of immune senescence and it was therefore noteworthy that spike-specific cellular responses were of similar magnitude within both staff and residents. Indeed, cellular responses against nucleocapsid protein were increased in donors aged over 40 years and therefore mirrored the increased profile of humoral immunity against this protein in relation to age. Functional activity was demonstrated by IFN-γ production within the ELISpot assay whilst IL-2 and IL-10 were also produced by virus-specific T cells. IL-2 production is a characteristic feature of the SARS-CoV-2 cellular response and suggests that the SARS-CoV-2-specific cellular response resides within a moderately differentiated effector pool but is not driven to terminal differentiation. Less is known regarding the role of the immunosuppressive cytokine IL-10. Cellular responses against both the spike and nucleocapsid proteins were stable over the 4 months of follow up and again concur with data from younger donors(11, 21).

Phenotyping of virus-specific CD4+ T cells indicated a moderately differentiated CD27+CD28+ phenotype which is consistent with the predominant IL-2 cytokine phenotype. It was notable that expression of the HLA-DR activation marker was markedly increased on T cells from LTCF residents and this increased further over time. This suggests ongoing chronic activation of virus-specific cells in older people, potentially due to increased retention of viral antigen, and this will be important to assess in longer term follow up.

Finally, we also measured serum concentrations of several cytokines across the life course and related these to SARS-CoV-2 serostatus. An increase in IL-6 and TNF-α concentration with age became apparent and is widely recognised as a marker of ‘inflammaging’(22), a phenomenon that is likely to be pronounced within the frail population within residential homes. Importantly, however, viral serostatus did not have any influence on the magnitude of this trajectory indicating that history of previous infection does not accelerate inflammation within this age group.

Vaccination has emerged as a highly effective approach to elicit protection against severe Covid-19 infection(7). As such it is important to interpret these findings in relation to vaccine responses in this population. Previous natural infection with SARS-CoV-2 strongly enhances spike-specific immune responses following vaccination and our findings likely underlie the 23-fold elevation of peak antibody responses after single vaccination in SARS-CoV-2-seropositive LTCF residents(23). These data indicate that short term vaccine responses in LTCF residents are likely to be comparable to those of younger people. It will be critical to assess if the comparable maintenance of immunity after natural infection translates into a similar profile after vaccination. This information will be critical in guiding the potential need for booster vaccination.

Potential limitations of our study include the fact that all seropositive donors clearly represent survivors of acute SARS-CoV-2 infection and, as mortality rates were high within the LTCF resident age group, there may have been potential selection bias for donors with the most effective underlying immune function. Additionally, we did not have access to the exact time or severity of primary infection or history of patient co-morbidities.

These findings indicate that immune responses to SARS-CoV-2 infection within the elderly and potentially frail resident LTCF population appear robust and comparable to those seen within younger people. These findings are consistent with similar levels of protection against reinfection within this cohort and provide insight into the potent influence of previous infection on the magnitude of the immune response to Covid-19 vaccination. It will now be important to assess how infection status acts to support the longeity of vaccine-induced immune responses and if this should be used as a determinant of the need for vaccine boosters.

## Methods

### Sample Collection

The VIVALDI study (ISRCTN14447421) (ongoing) is a prospective cohort study, which was set up in May 2020 to investigate SARS-CoV-2 transmission, infection outcomes, and immunity in residents and staff in LTCFs in England that provide residential and/or nursing care for adults aged 65 years and over (https://wellcomeopenresearch.org/articles/5-232/v2; (24)). Eligible LTCFs were identified by the Care Provider’s Senior Management Team, or by the National Institute for Health Research (NIHR) Clinical Research Network. Pseudonymised clinical (vaccination status, PCR test results) and demographic (age, sex, staff member versus resident) data were retrieved for staff and residents from participating LTCFs through national surveillance systems. All participants provided written informed consent for blood sample collection or if residents lacked the capacity to consent, a personal or nominated consultee was identified to act on their behalf.

Blood sampling was offered to participants at three-time points in June-July, August-September and October-November 2020. This time period was before the national vaccine rollout and therefore all data assess the immune response to natural infection alone. Anti-coagulated (EDTA) sample was sent to the University of Birmingham and serum tube to The Doctors Laboratory for anti-nucleocapsid IgG (N) testing. Ethical approval for this study was obtained from the South Central - Hampshire B Research Ethics Committee, REC Ref: 20/SC/0238.

### Data linkage

Abbott antibody test results were submitted to the COVID-19 datastore (https://data.england.nhs.uk/covid-19/) and linked to routinely held data on age, sex, LTCF, role (staff or resident) obtained through the national SARS-CoV-2 testing programme and to vaccination status (date and vaccine type) derived from the National Immunisations Management System (NIMS). These records were linked using a common identifier based on the individuals’ NHS number. Individual-level records were further linked to each LTCF using the unique Care Quality Commission location ID (CQC-ID), allocated by the Care Quality Commission who regulate all providers of health and social care in the UK.

### Inclusion criteria

Both staff and residents were eligible for inclusion if it was possible to link them to a pseudo-identifier in the COVID-19 Datastore. Only those that had been infected in the first wave of the pandemic in the UK or those that had no evidence of being infected were included in this sub-cohort. Samples from LTCFs that had experienced outbreaks in the first wave were chosen for seropositive samples and LTCFs with no outbreaks were chosen for the seronegative samples. Participants who seroconverted during the 4-month period of follow up were not included in the analysis. Due to limited PCR testing in the first wave of the pandemic, it was not possible to determine the date of primary infection with SARS-CoV-2. 21st April 2020 was the peak of the first wave for SARS-CoV-2 infection within LTCFs. Past infection with SARS-CoV-2 was defined based on results of Abbott antibody and MSD quantitative test using thresholds and methods outlined below.

### Sample Preparation

Samples were processed within 24 hours of reception at the University of Birmingham. Blood was spun at 300g for 5 minutes. Plasma was removed and spun at 500g for 10 minutes prior to storage at -80°C. The remaining blood was separated using a SepMate (Stemcell) density centrifugation tube. The resulting PBMC layer was washed twice with RPMI and frozen in freezing media (10% DMSO 90% FBS).

### Serological analysis of SARS-CoV-2-specific immune response

Quantitative IgG antibody titres were measured against trimeric Spike (S) protein. Multiplex MSD Assays were performed as per manufacturer instructions (Lot number K0081681). Briefly, 96-well plates were blocked. Following washing, samples were diluted at 1:5000 in diluent and added to the wells with the reference standard and internal controls. After incubation, plates were washed and anti-IgG detection antibodies were added. Plates were washed and were immediately read using a MESOTM QuickPlex SQ 120 system. Data was generated by Methodological Mind software and analysed with MSD Discovery Workbench (v4.0) software. Presented data were adjusted for any sample dilutions

For assessment of serological response against the SARS-CoV-2 nucleocapsid, blood samples were tested for the presence of IgG antibodies specific for nucleocapsid (N) protein using the Abbott ARCHITECT system (Abbott, Maidenhead, UK), a semi-quantitative chemiluminescent microparticle immunoassay. The assay was kindly performed by The Doctors Laboratory (London, UK). An index value cut-off of 0.8 was used to classify samples as antibody-positive (≥ 0.8)(25, 26).

### Quantitative inhibition of ACE-2 binding to SARS-CoV-2 spike

Quantitative inhibition of ACE-2 binding to trimeric SARS-CoV-2 Spike protein from variants of concern were measured using the MSD V-PLEX COVID-19 ACE2 Neutralization Kit (SARS-CoV-2 Plate 7) following manufacturer’s instructions (Lot number K0081681). Briefly, 96-well plates were blocked. Following washing, samples diluted 1:10 in the diluent, as well as reference standards, were added to the plate. After incubation, SULFO-TAG Human ACE-2 Protein detection protein was added to the plate and incubated for 1 hour. Plates were washed prior to reading immediately using a MESO ™ QuickPlex SQ 120 system. Data was generated by Methodological Mind software and analysed with MSD Discovery Workbench (v4.0) software. Presented data were adjusted for any sample dilutions.

### Quantification of SARS-CoV-2-specific cellular responses by ELISpot analysis

Pepmix pools containing 15-mer peptides overlapping by 10aa from either SARS-CoV-2 Spike S1 or S2, Nucleocapsid, Envelope and Membrane protein domains were purchased from Alta Biosciences (University of Birmingham, UK). T cell responses of post-vaccination samples to the above peptide mixes were determined using a Human IFN-γ ELISpot PRO kit (Mabtech, Sweden). Isolated PBMC were thawed and rested overnight prior to assay in R10 (RPMI + 10% FBS + Pen/Strep). 2-3×10□ PBMC were stimulated in duplicate with peptide mixes at 1ug/ml per peptide, anti-CD3 and CEFX cell stimulation mix (JPT) as a positive control, or DMSO as a negative control for 16-18 hours. Supernatants were harvested and stored at -80°C. Following the development of plates per the manufacturer’s instructions, the plates were read using the BioSys Bioreader 5000. Mean spot counts in DMSO treated negative control wells were deducted from the means to generate normalised spot counts for all other treated wells. Cut off values were previously determined (Zuo, et al. 2020).

Cytokine concentrations within ELISpot supernatants were assayed using a LEGENDplex™ Human Th Panel (BioLegend) according to manufacturer’s instructions. Data was analysed using the LEGENDplex™ Data Analysis Software Suite (BioLegend).

### Assessment of plasma cytokine levels

Multiplex assays to quantitatively measure levels of ten different cytokines (IFN-γ, IL-1β, IL-2, IL-4, IL-6, IL-8, IL-10, IL-12p70, IL-13 and TNF-□) within plasma were performed using the MSD V-PLEX Proinflammatory Panel 1 (human) kit according to manufacturer’s instructions (Lot number K0081665). Briefly, 96-well plates were washed prior to addition of serum diluted 1:4 in diluent and calibrators. Following incubation, plates were washed and detection antibodies were added. Plates were washed and read immediately using a MESO ™ QuickPlex SQ 120 system. Data was generated by Methodological Mind software and analysed with MSD Discovery Workbench (v4.0) software. Presented data was adjusted for any sample dilutions.

### Cell surface staining

Cryopreserved PBMC were thawed and rested for at least 6 hours in filtered R10. 10^6^+ cells were then stimulated with a combined SARS-CoV-2 Spike S1 and S2 peptide pool or a Nucleocapsid, Envelope and Membrane protein peptide pool at final concentration of 1μg/ml per peptide, or DMSO for an unstimulated control. Purified anti-CD40 antibody was added to the cell suspension at final concentration 1μg/ml. Cells were incubated at 37°C overnight for 16 hours. Following stimulation, cells were washed (PBS+5% BSA+1% EDTA), Brilliant Stain Buffer (BD) was added and cells were surface stained at 4°C for 30 minutes (supplementary table 2). Cells were washed and run on a BD Symphony A3 flow cytometer (BD Biosciences) and analysis was carried out using FlowJo v10.7.1 and Cytobank 2021.

### Statistical analysis

All data were checked for normality using the Kolmogorov-Smirnov test. For comparative analysis of 2 groups a Mann Whitney test was applied. Comparative analysis with 3 or more groups a Kruskal-Wallis test was used and for multiple comparisons uncorrected Dunn’s test was used for non parametric data. Spearman’s rank correlation coefficients were calculated and tested for correlations. Data analysis was performed in Graph Pad Prism V.9.1.0 (216).

## Data Availability

De-identified test results and limited meta-data will be made available for use by researchers in future studies, subject to appropriate research ethical approvals, once the VIVALDI study cohort has been finalised. These datasets will be accessible via the Health Data Research UK Gateway https://www.healthdatagateway.org/.

## Acknowledgements

The authors would like to thank the staff and residents in the LTCFs that participated in this study, and Mark Marshall at NHS England who pseudonymised the electronic health records. This report is independent research funded by the Department of Health and Social Care (COVID-19 surveillance studies). AH is supported by Health Data Research UK (HDR-UK; grant no LOND1), which is funded by the UK Medical Research Council, Engineering and Physical Sciences Research Council, Economic and Social Research Council, Department of Health and Social Care (England), Chief Scientist Office of the Scottish Government Health and Social Care Directorates, Health and Social Care Research and Development Division (Welsh Government), Public Health Agency (Northern Ireland), British Heart Foundation, and Wellcome Trust. LS is funded by a National Institute for Health Research (NIHR) Clinician Scientist Award (CS-2016-007). MK is funded by a Wellcome Trust Clinical PhD Fellowship (222907/Z/21/Z). The views expressed in this publication are those of the authors and not necessarily those of the NHS, Public Health England, or the Department of Health and Social Care.

## Contributors

Study conceptualisation: LS, AC, AH, MK, GT, PM;

Method conceptualization: GT, AD, PM, HP

Project administration: MK, BA, CF, RB, GT, CB, UA, SH, AJ, DB, MB, MA, ES;

Data curation and validation: GT, TL, MB, ES, PS, NK,;

Funding: LS, AH, ACm, PM;

Writing (original draft): GT, PM, TL, MB;

Writing (review and editing): All authors. GT, TL, NK, PS, MB, AD and PM had full access to the data in the study. PM and GT have shared responsibility for the decision to submit for publication.

