## Supplement for "Robust SARS-CoV-2-specific and heterologous immune responses after natural infection in elderly residents of Long-Term Care Facilities"

| **Spike** | **<40 Col1** | **<40 Col2** | **<40 Col3** | **40-64 Col1** | **40-65 Col2** | **40-64 Col3** | **65-84 Col1** | **65-84 Col2** | **65-84 Col3** | **>85 Col1** | **>85 Col2** | **>85 Col3** |
| --- | --- | --- | --- | --- | --- | --- | --- | --- | --- | --- | --- | --- |
| **Number of values** | 19 | 19 | 19 | 67 | 67 | 67 | 42 | 42 | 42 | 35 | 35 | 35 |
| **Minimum** | 191.1 | 86.55 | 99.61 | 388.1 | 31.37 | 31.75 | 62.46 | 17.45 | 49.23 | 64.18 | 32.92 | 49.65 |
| **25% Percentile** | 1848 | 3773 | 2982 | 3164 | 3788 | 3251 | 2913 | 2531 | 2185 | 1950 | 3057 | 2485 |
| **Median** | 5044 | 7079 | 5995 | 6670 | 7615 | 6539 | 6102 | 6519 | 8136 | 9788 | 12798 | 10419 |
| **75% Percentile** | 13568 | 16999 | 15834 | 18386 | 16959 | 14119 | 19060 | 18110 | 20998 | 22645 | 28968 | 23795 |
| **Maximum** | 105561 | 113477 | 82380 | 189014 | 194872 | 147131 | 146310 | 187957 | 359073 | 254745 | 249242 | 203003 |
| **Range** | 105370 | 113390 | 82281 | 188626 | 194840 | 147100 | 146248 | 187940 | 359024 | 254681 | 249209 | 202953 |
| **Mean** | 12158 | 14914 | 13327 | 13821 | 13831 | 12391 | 20570 | 23185 | 28336 | 23279 | 27096 | 21867 |
| **Std. Deviation** | 23334 | 25377 | 20470 | 24836 | 24851 | 19930 | 33146 | 39691 | 62677 | 44518 | 46553 | 39112 |
| **Std. Error of Mean** | 5353 | 5822 | 4696 | 3034 | 3036 | 2435 | 5115 | 6125 | 9671 | 7525 | 7869 | 6611 |
| **Geometric mean** | 4791 | 5712 | 5255 | 6759 | 6225 | 5687 | 6303 | 5921 | 6618 | 6122 | 6742 | 5794 |
| **Geometric SD factor** | 4.217 | 5.238 | 4.979 | 3.359 | 4.603 | 4.49 | 5.944 | 8.106 | 6.992 | 8.182 | 9.819 | 7.953 |
| **RBD** |  |  |  |  |  |  |  |  |  |  |  |  |
| **Minimum** | 200.8 | 70.01 | 88.52 | 132.9 | 27.92 | 42.08 | 57.27 | 61.53 | 71.9 | 66.7 | 21.24 | 36.81 |
| **25% Percentile** | 457.4 | 654.7 | 562.6 | 1243 | 1147 | 997.6 | 1011 | 752.1 | 664.9 | 527.9 | 681 | 656.5 |
| **Median** | 1588 | 2041 | 1456 | 2446 | 2675 | 2353 | 2181 | 2385 | 2464 | 3356 | 3925 | 2839 |
| **75% Percentile** | 5012 | 4817 | 4178 | 5446 | 6393 | 5156 | 6893 | 6121 | 6279 | 9162 | 11288 | 8220 |
| **Maximum** | 7889 | 10209 | 15719 | 62434 | 65125 | 51954 | 98386 | 79251 | 79291 | 63940 | 58235 | 50117 |
| **Range** | 7689 | 10139 | 15631 | 62301 | 65097 | 51912 | 98328 | 79190 | 79220 | 63873 | 58214 | 50080 |
| **Mean** | 2557 | 3398 | 3250 | 5092 | 5021 | 4435 | 8515 | 9385 | 7964 | 7220 | 9113 | 7677 |
| **Std. Deviation** | 2405 | 3261 | 3838 | 8693 | 8657 | 7236 | 17850 | 19050 | 15713 | 11571 | 13766 | 12123 |
| **Std. Error of Mean** | 551.8 | 748.2 | 880.5 | 1062 | 1058 | 884 | 2754 | 2939 | 2454 | 1956 | 2327 | 2049 |
| **Geometric mean** | 1434 | 1823 | 1677 | 2443 | 2328 | 2085 | 2449 | 2405 | 2251 | 2421 | 2842 | 2474 |
| **Geometric SD factor** | 3.38 | 3.865 | 3.719 | 3.431 | 3.969 | 3.822 | 5.009 | 5.647 | 5.29 | 5.92 | 6.588 | 5.752 |
| **Nucleocapsid** |  |  |  |  |  |  |  |  |  |  |  |  |
| **Minimum** | 454.8 | 152.1 | 143.7 | 90.23 | 79.61 | 70.33 | 25.22 | 41.99 | 41.21 | 25.22 | 41.99 | 41.21 |
| **25% Percentile** | 3353 | 3744 | 2230 | 6571 | 5706 | 3808 | 4679 | 3138 | 2964 | 2532 | 2406 | 1920 |
| **Median** | 12522 | 13399 | 8648 | 13962 | 12208 | 7398 | 24697 | 14299 | 11819 | 28699 | 27154 | 15553 |
| **75% Percentile** | 32511 | 40948 | 24910 | 32553 | 24819 | 18818 | 44674 | 44235 | 32908 | 55249 | 57078 | 31568 |
| **Maximum** | 81511 | 84452 | 51809 | 134245 | 164144 | 115855 | 194471 | 170952 | 110074 | 308046 | 359406 | 333983 |
| **Range** | 81056 | 84300 | 51665 | 134155 | 164064 | 115785 | 194445 | 170910 | 110032 | 308021 | 359364 | 333942 |
| **Mean** | 23219 | 22845 | 15281 | 26860 | 24302 | 15633 | 31968 | 28207 | 22497 | 55101 | 51040 | 33431 |
| **Std. Deviation** | 26755 | 26447 | 17037 | 31084 | 32163 | 20881 | 38607 | 35695 | 26623 | 80737 | 77902 | 62637 |
| **Std. Error of Mean** | 6138 | 6067 | 3909 | 3798 | 3929 | 2551 | 5957 | 5508 | 4158 | 13647 | 13168 | 10588 |
| **Geometric mean** | 10019 | 8336 | 5943 | 13503 | 10339 | 6875 | 13551 | 10726 | 8381 | 14921 | 13503 | 8603 |
| **Geometric SD factor** | 4.838 | 6.336 | 5.949 | 3.832 | 4.965 | 4.56 | 5.369 | 5.619 | 5.812 | 7.95 | 8.174 | 7.242 |

**Supplementary table 1: Median, mean, and geometric mean values of spike, RBD and nucleocapsid IgG response in seropositive donors.**

| **Antigen** | **Clone** | **Fluorophore** | **Supplier** | **Code** |
| --- | --- | --- | --- | --- |
| BD Horizon Fixable Viability Dye | - | FVS575V | BD | 565694 |
| CD14 | M5E2 | BV650 | Biolegend | 301836 |
| CD19 | HIB19 | BV650 | Biolegend | 302238 |
| CD3 | SK7 | AF700 | Biolegend | 344822 |
| CD8 | SK1 | BUV805 | BD | 612889 |
| CD4 | SK3 | BUV496 | BD | 612936 |
| CD27 | L128 | BUV563 | BD | 748705 |
| CD25 | 2A3 | BUV615 | BD | 612996 |
| CD127 | HIL-7R-M21 | BUV737 | BD | 612794 |
| CD45RA | HI100 | BV480 | BD | 566114 |
| CCR7 | 2-L1-A | APC-Cy7 | Biolegend | 353212 |
| CD69 | FN50 | BV711 | BD | 563836 |
| HLA-DR | G46-6 | BV786 | BD | 564041 |
| TCRgd | 11F2 | BB700 | BD | 745944 |
| CD56 | B159 | PE-Cy5 | BD | 555517 |
| CD28 | CD28.2 | BUV661 | BD | 741635 |
| CD39 | TU66 | A1 | Biolegend | 328224 |
| CD95 | DX2 | PE-Cy7 | Biolegend | 305622 |
| CD154 | 24-31 | APC | Biolegend | 310810 |
| CD137 | 4B4-1 | PE | Biolegend | 309804 |
| Purified anti-CD40 | HB14 | **-** | Biolegend | 313020 |

**Supplementary table 2: Cell surface staining panel (AIM)**

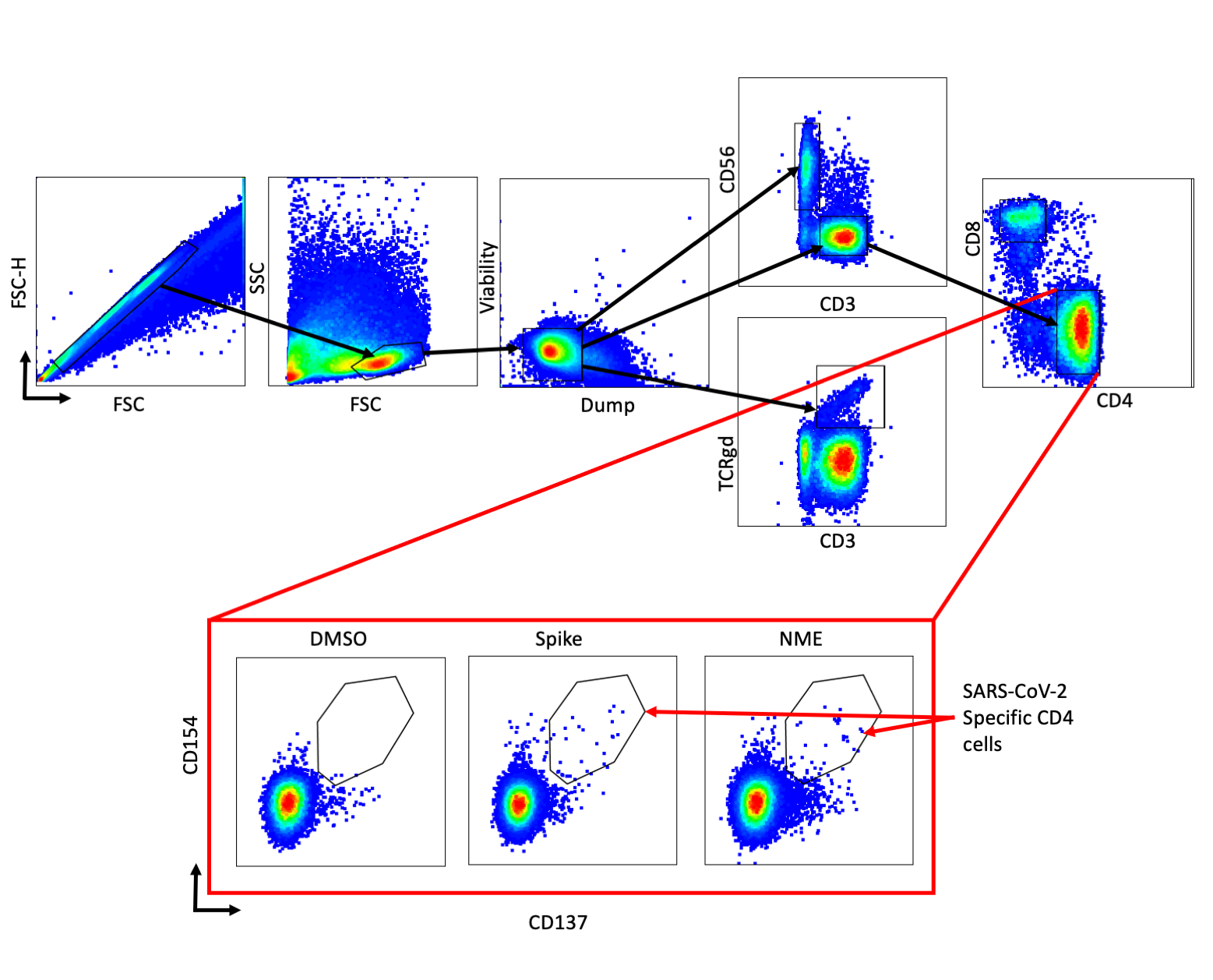

**Supplementary figure 1: AIM gating strategy to isolate SARS-CoV-2 specific CD4 cells.**

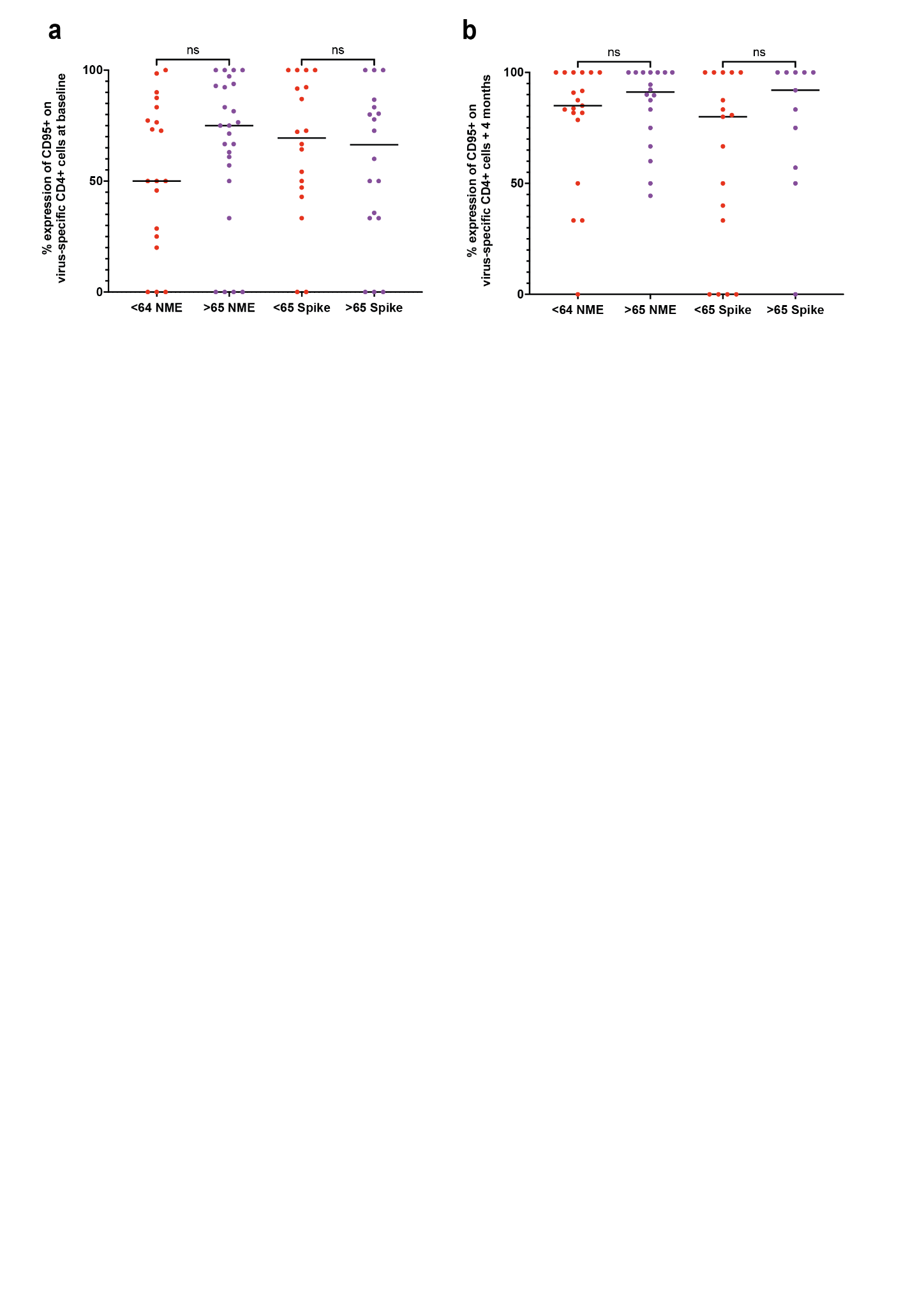

**Supplementary figure 2: Percentage of CD95+ on virus specific CD+ cells at baseline and + 4 months.**

a/b) Quantification of CD95+ on CD4+ virus-specific cells after stimulation with N/M/E or spike peptides. Red dots are donors aged <65 years and purple dots are donors aged >65 years.
